# Interstitial Cystitis: a phenotype and rare variant exome sequencing study

**DOI:** 10.1101/2025.02.16.25322147

**Authors:** Joshua E. Motelow, Ayan Malakar, Sarath Babu Krishna Murthy, Miguel Verbitsky, Atlas Kahn, Elicia Estrella, Louis Kunkel, Madelyn Wiesenhahn, Jaimee Becket, Natasha Harris, Richard Lee, Rosalyn Adam, Krzysztof Kiryluk, Ali G. Gharavi, Catherine A. Brownstein

**Author notes:** Corresponding author. (J.E.M.); (C.A.B.).

## Abstract

Interstitial cystitis/bladder pain syndrome (IC/BPS) is a poorly understood and underdiagnosed syndrome of chronic bladder/pelvic pain with urinary frequency and urgency. Though IC/BPS can be hereditary, little is known of its genetic etiology. Using the eMERGE data, we confirmed known phenotypic associations such as gastroesophageal reflux disease and irritable bowel syndrome and detected new associations, including osteoarthrosis/osteoarthritis and Barrett’s esophagus. An exome wide ultra-rare variants analysis in 348 IC/BPS and 11,981 controls extended the previously reported association with *ATP2C1* and *ATP2A2,* implicated in Mendelian desquamating skin disorders, but did not provide evidence for other previously proposed pathogenic pathways such as bladder development, nociception or inflammation. Pathway analysis detected new associations with “anaphase-promoting complex-dependent catabolic process”, the “regulation of MAPK cascade” and “integrin binding”. These findings suggest perturbations in biological networks for epithelial integrity and cell cycle progression in IC/BPS pathogenesis, and provide a roadmap for its future investigation.

## Introduction

Interstitial cystitis/bladder pain syndrome (IC/BPS) is a disorder of chronic pain including symptoms of urinary frequency, urgency, and bladder, suprapubic, and lower back discomfort or pain (*1, 2*). Diagnostic criteria have varied over time, but prevalence estimates in the United States suggest 2.7%-6.5% of adult women and 1.9%-4.2% of men have IC/BPS (*1, 3–5*). IC/BPS is likely underdiagnosed, and there is often a long diagnostic odyssey due to the lack of biomarkers or known etiology (*3, 4, 6*).

The pathophysiology of IC/BPS is incompletely understood (*7–15*). On cystoscopy, up to 10-20% of affected individuals may have the characteristic Hunner lesions/ulcers that are pathognomonic for IC/BPS, but this finding has low sensitivity for IC/BPS as a whole (*16–18*). Multiple studies suggest a genetic predisposition. There is a greater concordance of IC/BPS diagnoses in monozygotic than dizygotic twins and in 1^st^ degree relatives than in the general population (*19, 20*). Bladder pain scores in monozygotic twins are correlated (*21*). Single nucleotide polymorphisms (SNPs) previously associated with conditions often co-morbid with IC/BPS were more prevalent in individuals with IC/BPS than in controls (*22*). Finally, our group has previously identified Mendelian disorders within an IC/BPS cohort (*23*).

Individuals with IC/BPS are affected with other comorbidities at increased rates. Previous literature supports an association with nociplastic pain disorders (e.g., irritable bowel syndrome, fibromyalgia, and encephalomyelitis/chronic fatigue syndrome [ME/CFS]), anxiety, and depression (*4, 24–26*). Other previously noted associations include reflux esophagitis (*27*), urinary calculus (*28*), allergic rhinitis (*29*), and vomiting (*30*).

This study, to our knowledge, is the largest exome sequencing (ES) study of individuals with IC/BPS to date. Using a new phenotype database, we reaffirm the association of previously identified co-morbid phenotypes while identifying new associations. We have identified genes with multiple carriers of ultra-rare deleterious variants for which no control carriers were identified. We used genes with excess ultra-rare damaging variants in individuals with IC/BPS to interrogate gene pathways. We identified significant associations in the Gene Ontology (GO) biological process pathway “anaphase-promoting complex-dependent catabolic process” (GO:0031145) and identified a significant association with the “transport of small molecules” (R-HSA-382551) Reactome pathway (*31–35*). This study strengthens the prior association of IC/BPS with deleterious variants in genes with known Mendelian disease associations and takes another step toward unraveling the etiology of this debilitating disorder.

## Results

### IC case-control cohort demographics

We combined exome sequencing from two cohorts (“BCH cohort” and “MaGIC cohort”, see Materials and Methods) of individuals with IC/BPS (Table 1) for case-control gene, variant, and gene-set analysis. The BCH cohort has been previously described (*23*). It included exome sequencing from 381 individuals (319 female) including probands, affected family members and unaffected family members. 283 individuals were probands. The MaGIC cohort included 100 individuals selected for sequencing from a larger cohort (see Materials and Methods) (*36*). All individuals in the MaGIC cohort were female probands. In the combined cohort of analyzed individuals, 43 individuals were also diagnosed with anxiety (1 in MaGIC), 88 individuals with migraine (48 in MaGIC), 6 with Hunner lesions (0 in MaGIC), and 55 with irritable bowel syndrome (IBS, 55 in MaGIC).

**Table 1.** Subject demographics. Demographics of probands across two cohorts. See Materials and Methods for cohort descriptions. Ancestry predicted from exome sequencing data (see Methods). No age of diagnosis data available from MaGIC cohort. For BCH cohort, age of diagnosis available for 204 (190 female) of 250 individuals.

|  | BCH cohort |  |  | MaGIC cohort |
| --- | --- | --- | --- | --- |
| Geographic Ancestry | Total (% of population) | Females | Males | Total (100% females) |
| Latino | 6 (1.7%) | 6 (1.7%) | 0 (0%) | 3 (0.9%) |
| European | 221 (92.4%) | 202 (58%) | 19 (5.5%) | 64 (18.4%) |
| East Asian | 2 (0.6%) | 2 (0.6%) | 0 (0%) | 0 (0%) |
| Middle Eastern | 13 (3.7%) | 11 (3.2%) | 2 (0.6%) | 6 (1.7%) |
| Admixed | 8 (2.3%) | 6 (1.7%) | 2 (0.6%) | 25 (7.1%) |
| Total | 250 | 227 | 23 | 98 |
| Age of Diagnosis | BCH cohort | Females | Males | MaGIC cohort |
| Min, 1<sup>st</sup> Quartile, Median, Mean, 3<sup>rd</sup> Quartile, Max | 8.0, 30.0, 43.0, 41.8, 52.25 | 10.0, 30.0, 43.0, 41.9, 52.75, 75 | 8.0, 26.25, 39.0, 39.6, 48.0, 74 | Unavailable |

Though some families had multiple potentially affected family members, affected family members may not have undergone the same rigorous evaluation for inclusion and exclusion criteria. Therefore, our collapsing analysis only included probands who were screened for inclusion to either of the two cohorts (BCH and MaGIC). 348 individuals with IC/BPS (325 females) and 11,669 controls (6,596 females) were included in exome analysis.

### Phenome-wide co-morbidity analysis

Prior epidemiological analyses identified many comorbidities with IC/BPS including fibromyalgia, irritable bowel syndrome, and anxiety (*4, 24–26, 37*). We performed a phenome-wide comorbidity analysis across 1,816 phecodes in the Electronic Medical Records and Genomics (eMERGE) network dataset (*38, 39*) (Fig. 1), comparing 360 individuals (316 female) with IC/BPS to 99,249 individuals (53,224 female) without IC/BPS (Data S1). We validated prior comorbidities, including dysuria (odds ratio [OR] 12.3, *P* = 5.1e-23) (*1, 2*), polyuria (OR 77.2, *P* = 2.1e-49) (*1, 2*), irritable bowel syndrome (OR 6.5, *P* = 6.3e-7) (*40*), esophagitis (OR 2.7, *P* = 8.3e-6) (*27*), gastroesophageal reflux disease (OR 2.8, *P* = 1.3e-5) (*27*), and allergic rhinitis (OR 3.8, *P* = 1.1e-6) (*41*). Novel associations were identified such as Barrett’s esophagus (OR 11, *P* = 1.9e-5) and osteoarthrosis/osteoarthritis (OR 3.2, *P* = 1.1e-5). These provide new candidate biological pathways to investigate for IC/BPS. Prior phenotypic associations such as chronic pain (OR 2.8, *P* = 0.00030) (*24*) and anxiety (OR 2.2, *P* = 0.0010) (*42*), among others, were suggestive.

**Fig 1.**
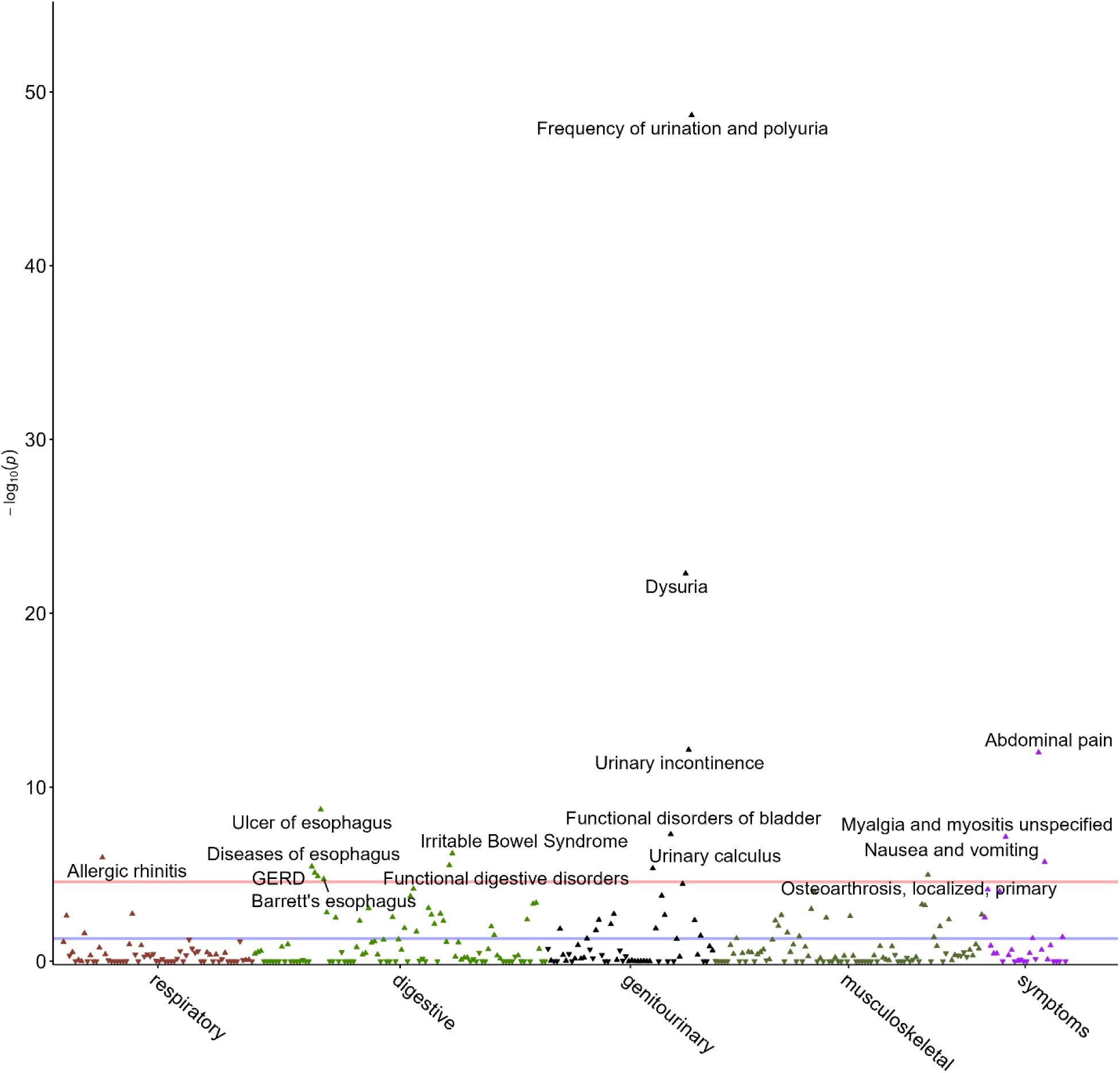
Phenotype analysis using eMERGE. Comorbidity analysis identifying phecodes in individuals with IC/BPS in eMERGE compared 360 individuals with IC/BPS to 99,249 individuals without IC/BPS (see methods). The association of having IC/BPS and carrying each of 1,816 phecodes was assessed. Statistical association was tested with logistic regression (see Methods). Blue horizontal line indicates 0.05, the red horizonal line indicates 0.05 / 1,816. Phecodes with p values < 2.7e-5 are annotated. Only phenotype categories with significant associations are shown. Full data available in Data S9. Labels for non-specific phecodes beginning in “Other”, Symptoms”, or “Functional” have been removed to improve readability.

### Gene-based collapsing

We first explored gene-based collapsing to identify associations of case/control status with individual genes (*43–46*). Each gene-based collapsing model was defined by a qualifying variant (Table S1). We created models with both ultra-rare (absent from control databases) and rare (minor allele frequency [MAF] < 1%) qualifying variants. We created models for damaging effects (damaging missense and predicted loss-of-function) and synonymous (presumed control). All models showed limited genomic inflation (L < 1.1) suggesting good ancestry sub-structure matching (Fig. S1 – S2, Data S2 – S3).

In our ultra-rare and rare damaging models, no association exceeded study-wide significance (Fig. 2, Fig. S3, Data S4 – S5). We highlighted seven genes (Table 2) in our ultra-rare functional model which had more than one case carrier of a qualifying variant (QV) and no control carriers (5,939 genes harbored multiple control carriers and no case carriers, data not shown). The top association was *DNAAF4* (MIM: 608706) for which 3 cases harbored ultra-rare functional variants and no controls harbored variants. *DNAAF4* is implicated as an autosomal dominant risk factor for dyslexia (*47*). Biallelic variants in *DNAAF4* cause ciliary dyskinesia (*48*). Among the remaining genes with multiple case carriers and no control carriers, the gene most consistent with the known prior disease associations with IC/BPS is *HTR7* (MIM: 182137) which encodes a serotonin receptor implicated in visceral hypersensitivity in irritable bowel syndrome (*49*). Other genes with at least 2 case carriers and no control carriers include *CALB2* (MIM: 114051), *SERPINF2* (MIM: 613168), *STX6* (MIM: 603944), *TBC1D26* (MIM: NA), and *MNT* (MIM: 603039) (*50–54*). None of the 13 individuals carrying variants in the above-mentioned genes had enrolled family members, and therefore we were unable to assess for segregation. Of the seven genes, two have a prior disease association but due to incomplete phenotyping information, we could not ascertain the presence or absence of related phenotypes in IC/BPS affected carriers. The more inclusive rare damaging model did not yield more compelling associations (Fig. S3, Data S4).

**Fig. 2.**
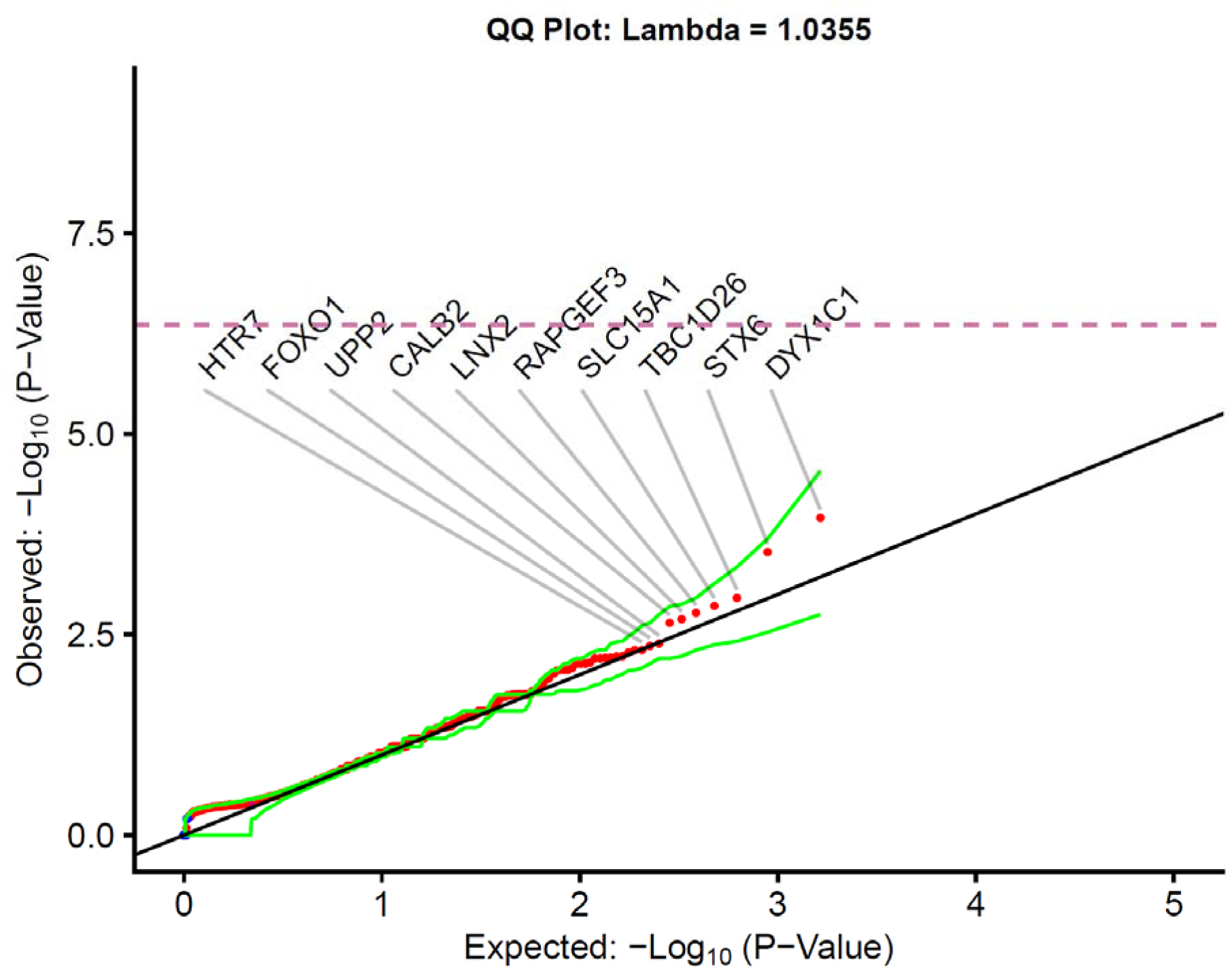
Ultra-Rare Damaging Missense and Loss-of-Function Model with Unrelated Probands. The quantile-quantile plots for the protein-coding genes with at least one case or control carrier of an ultra-rare predicted loss-of-function or predicted damaging missense variant. All variants are ultra-rare (i.e., allele frequency of less than 0.05% in internal case and control by cluster, absent in external reference cohorts, allele frequency < 0.002% in IGM database). P-values were generated from the exact two-sided Cochran-Mantel-Haenszel (CMH) test by gene by cluster to indicate a different carrier status of cases in comparison to controls. No gene achieved study-wide significance p < 5.4 x 10^-7^ after Bonferroni correction indicated by dashed line. Top ten case enriched genes are labeled. The green lines represent the 95% confidence interval. Red indicates case-enriched while blue indicates control enriched. Empiric confidence interval distributions created by permutations (n = 1,000). Included effects = stop-gain, frameshift, splice donor, splice acceptor, missense. All missense variants REVEL > 0.5. 331 individuals with IC/BPS compared to 6,516 controls.

**Table 2.**
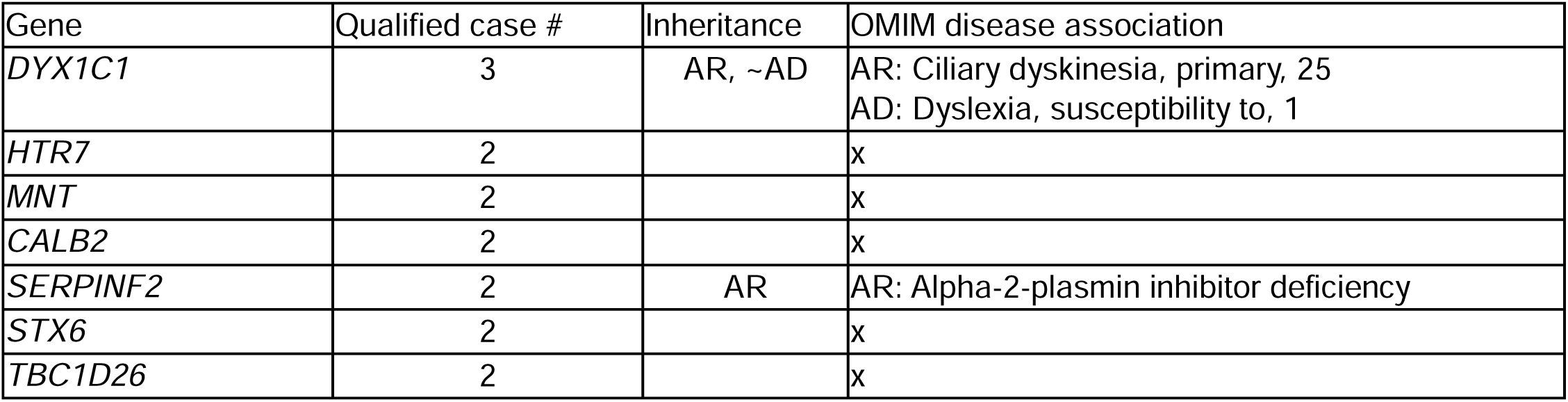
Candidate genes for ultra-rare variant risk for IC/BPS. Genes with multiple case hits but no control hits. Disease associations drawn from Online Mendelian Inheritance in Man (OMIM). “x” indicates no disease association in OMIM, AD = autosomal dominant, AR = autosomal recessive, ∼ = indicates gene associated with susceptibility to multifactorial disorders in OMIM.

### Gene set analyses

Using predefined gene sets drawn from novel (Fig. 1, Data S1) and previously associated phenotypes, we tested for enrichment of ultra-rare damaging variants in previously proposed pathogenic pathways (Table S2, Data S6, see Materials and Methods). We interrogated genes for developmental anomalies of the kidney and urinary tract genes (*55*), epithelial structure maintenance (GO:0010669), morphogenesis of an epithelial sheet (GO: 0002011), genes previously implicated in Mendelian diagnoses among individuals with IC/BPS (*23*), genes associated with desquamating dermatologic disorders (see Methods), GO biological function gene sets containing both *ATP2C1* (MIM: 604384) and *ATP2A2* (MIM: 108740), genes expressed in the bladder, genes important for bladder development (*56–70*), 4 genes recently implicated in animal models of IC/BPS (*71–74*), and genes encoded in GWAS loci for pain perception, depression, GERD, osteoarthrosis/osteoarthritis, allergic rhinitis, and irritable bowel syndrome (IBS) (*75–81*). This focused gene-set analysis did not provide any evidence supporting any of the previously proposed pathways. We only detected statistical evidence for the 5-gene set implicated in our prior analysis of Mendelian disorders in IC/BPS (*23*) (OR, 7.4; 95% CI, 1.6 – 26.4; unadjusted *P* = 0.0052). The association was supported both by the original and the new cohorts (MaGIC cohort; OR, 13.5; 95% CI, 1.4 – 68.2; unadjusted *P* = 0.014; BCH cohort; OR, 5.3; 95% CI, 0.6 – 25.7; unadjusted *P* = 0.071), and importantly, were driven by case variants in *ATP2C1* (3 cases) and *ATP2A2* (1 case), the most compelling candidate genes that emerged from the original analysis. Two IC/BPS affected proband carriers were identified in the previously analyzed BCH cohort and two additional carriers were found in the newly added MaGIC cohort. We found an additional missense variant (3-130678173-C-G) in *ATP2C1* that satisfied the strict requirements of our ultra-rare damaging model in one of the 17 individuals who were not included in the collapsing analysis because of inadequate representation of geographic ancestry (Fig. S4).

To perform a more comprehensive network-based pathway analysis, we applied the network-based heterogeneity clustering algorithm (NHC), a method designed to detect physiological homogeneity within a genetically heterogeneous dataset by testing enrichment within gene and protein networks constructed from interaction databases (Table 3, Data S7) (*82*). This approach yielded multiple significant gene clusters and pathway associations. The most significantly associated GO biological process pathway from the most significantly enriched gene cluster was “anaphase-promoting complex-dependent catabolic process” (Table 3, GO: 0031145, *P* = 6.2e-06) (*31, 32*), which includes *SMAD3* (MIM: 613795). The pathway has previously been implicated in cell cycle-cell proliferation and in multiple forms of cancers, including bladder cancer (*83, 84*). Additional noteworthy clusters included the *EGF* (MIM: 131530) and *PIK3R3* (MIM: 606076) genes (Data S7, Cluster 10), positive regulation of MAPK cascade (Table 3, GO: 0043410, *P* = 5.5e-06) and the integrin/focal adhesion networks (Data S7, Cluster 14). For an additional pathway analysis, we used fast preranked gene set enrichment analysis (fgsea) to detect pathways enriched in our ultra-rare variant analysis (*85, 86*). After accounting for multiple comparisons, the “transport of small molecules” pathway (GO:0006810, Reactome R-HSA-382551) remained significantly enriched (Figure 3, adjusted *P* = 0.042) (*31, 32, 35, 87*). This pathway includes *ATP2C1* and *ATP2A2* which drive the signal. Little is known about this pathway in the context of human disease (*88*).

**Fig 3.**
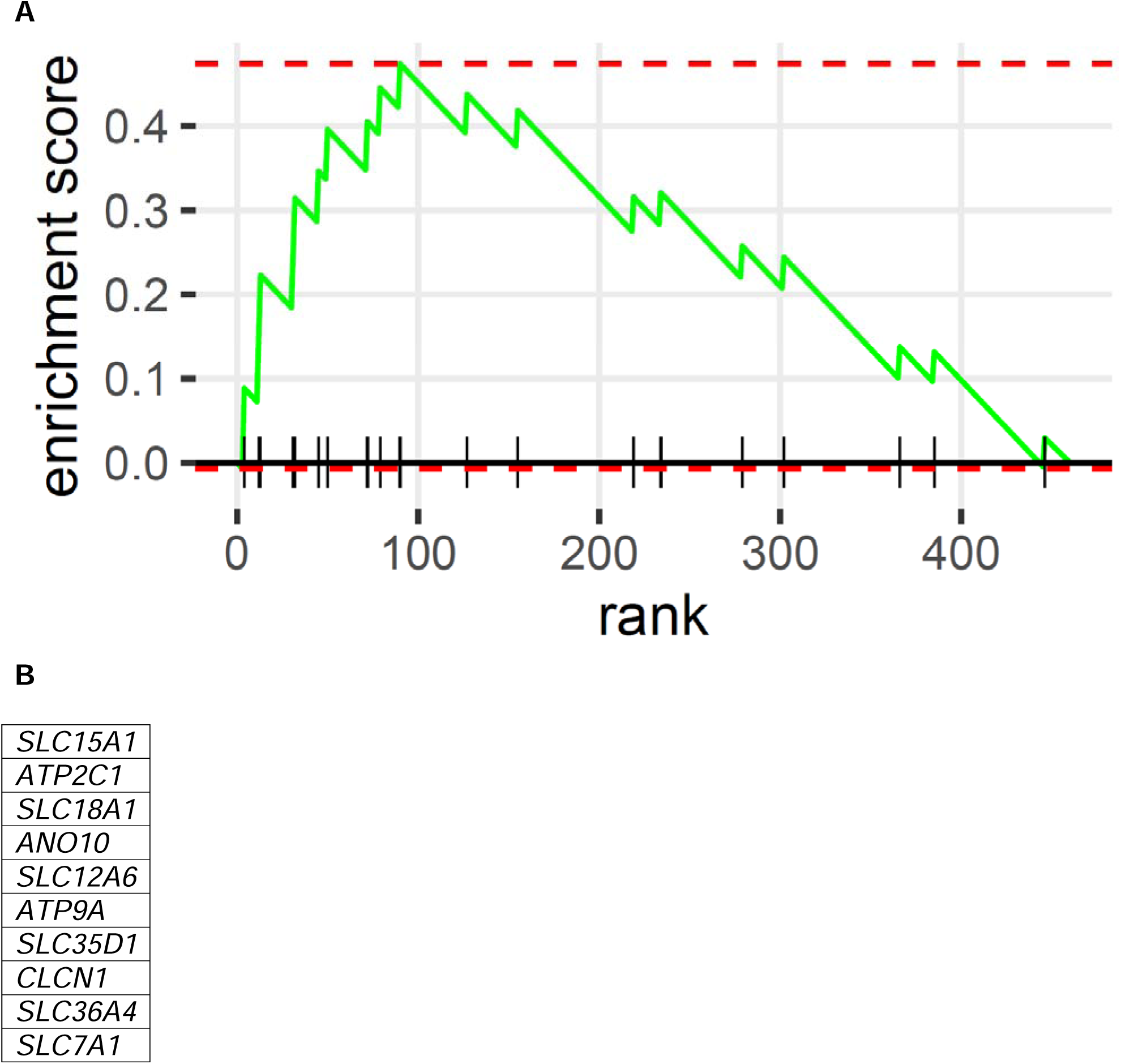
Gene set enrichment analysis. (A) The fGSEA plot of the only significantly associated hallmark gene set (“transport of small molecules”) using genes associated with IC/BPS in the ultra-rare functional collapsing model. The normalized enrichment score (NES) was 1.99 with an adjusted p value = 0.042. A p-value < 0.1 was considered the threshold for association for a gene to be included in fgsea analysis. The statistical threshold for pathway association was adjusted p < 0.05. Dashed red line indicates the positive enrichment score which indicates the maximum deviation from zero. The “normalize enrichment score” is obtained by normalizing the enrichment score to mean enrichment of random samples of the same size. P values vary slightly by run. (B) Leading edge genes which drive the enrichment of the “transport of small molecules” pathway. fGSEA = fast gene set enrichment analysis. R package “fgsea” was used for this analysis.

**Table 3.** Gene clusters and pathway enrichment detected by Network-based Heterogeneity Clustering (NHC). Damaging ultra-rare variant model used as input where sample-gene combination included if gene harbored a variant in the damaging ultra-rare model. Table includes enriched gene cluster which are associated with GO pathways detected by NHC. Full data in Data S7.

| **Genes** | **Cluster P-value** | **GO Pathway** |
| --- | --- | --- |
| *ANAPC1 ANAPC7 CDC20*<br>*CDC23 FOXO1 NEK2*<br>*PPARGC1A SKIL SMAD3*<br>*TRIM33* | 8.6e-08 | anaphase-promoting complex-dependent catabolic process (6.2e-06) |
| *EGF ERBB2 ERBB4 FGF10*<br>*FGFR1 FGFR4 IGF1R NRG2*<br>*PIK3R3* | 0.00043 | peptidyl-tyrosine modification (1.2e-07)<br>positive regulation of MAPK cascade<br>(5.5e-06)<br>transmembrane receptor protein tyrosine<br>kinase activity (3.2e-07)<br>Transmembrane receptor protein kinase<br>activity (1.2e-06) |
| *BCAR1 FLNC ITGA2 ITGA6*<br>*ITGA9 ITGB1 ITGB3 LAMA5*<br>*TLN2* | 0.0042 | integrin-mediated signaling pathway<br>(2.0e-10)<br>cell adhesion mediated by integrin (4.2e-06)<br>Integrin binding (1.1e-09) |

### Exome wide association study (ExWAS)

We performed a rare-variant (MAF < 1%) exome wide association study (ExWAS) (1). We performed a Cochran–Mantel–Haenszel (CMH) test on each site with a variant using the same clusters generated for the gene-based collapsing (Figure 4, Table S3, Data S8). The most significant association is a synonymous variant with a low Transcript-inferred Pathogenicity (TraP) score (0.41) in *SLC9A5* ([MIM: 600477] (1-48708137-C-A, 7 of 331 case carriers, 8 of 6,516 control carriers; OR, 16.2; 95% CI, 4.8 – 54.1; unadjusted *P* = 6.6 x 10e-6) (*89*). *SLC9A5* promotes tumor growth and cell motility, and it is part of the solute carrier protein superfamily implicated in our gene-set enrichment analysis (Figure 3B) (*90*). The second most significant association is a SNV in *CD68* [MIM: 153634] (17-7483627-A-G, 4 of 331 case carriers, 7 of 6,516 control carriers; OR, 38.6; 95% CI, 7.7 – 168.1; unadjusted *P* = 2.0 x 10-5). It is a synonymous variant with a TraP score of 0.141 (*89*). *CD68* encodes a scavenger receptor that has been implicated in pain syndromes (*91*).

**Fig 4.**
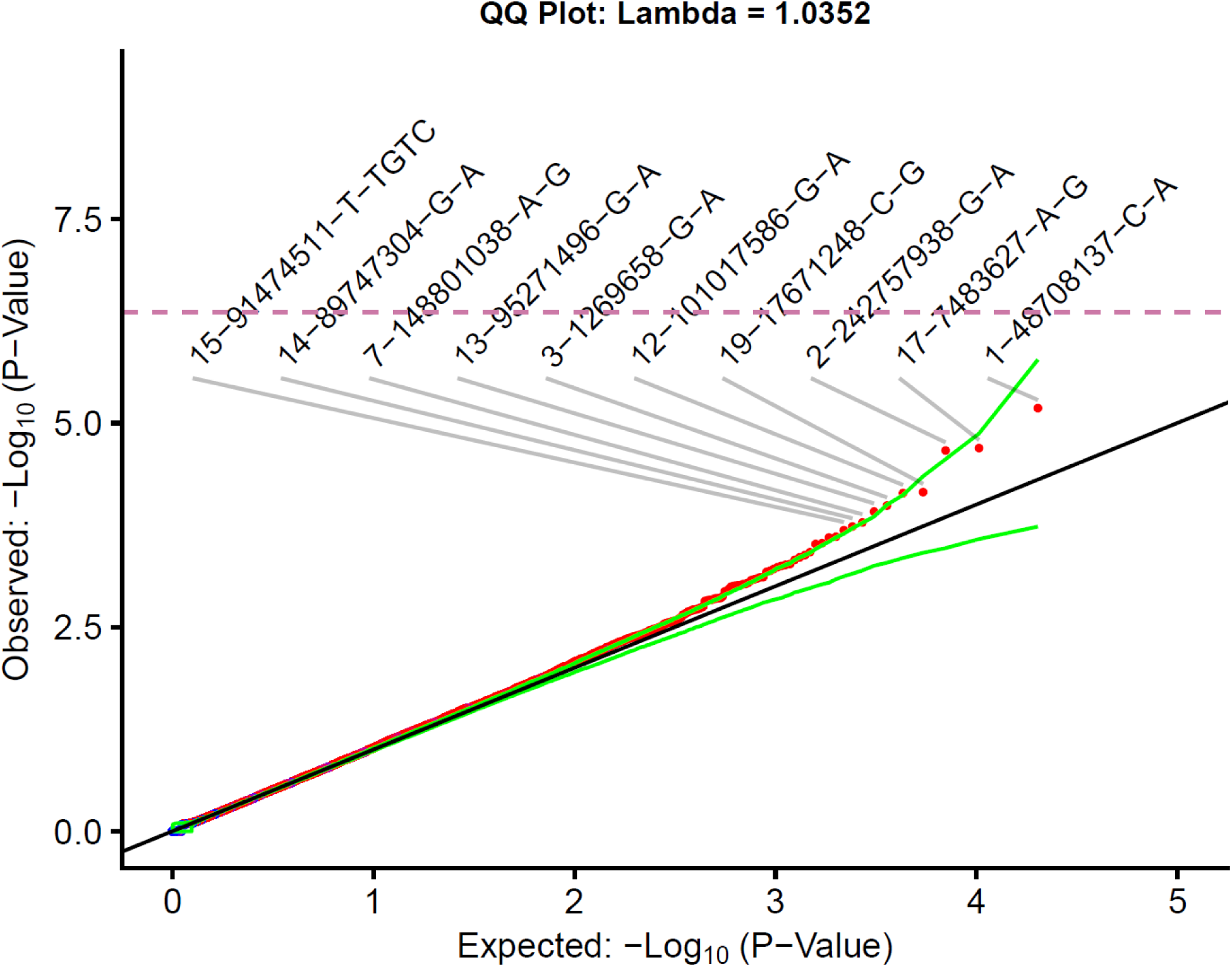
Rare variant exome wide association study. The quantile-quantile plots for variants in protein-coding genes with at least one case or control carrier. All variants had an allele count of at least 10 in eligible bases after coverage harmonization. P-values were generated from the exact two-sided Cochran-Mantel-Haenszel (CMH) test by gene by cluster to indicate a different carrier status of cases in comparison to controls. No variant achieved study-wide significance p < 5.4 x 10^-7^ after Bonferroni correction indicated by dashed line. Top ten case enriched variants are labeled. Red indicates case-enriched while blue indicates control enriched. The green lines represent the 95% confidence interval. Empiric confidence interval distributions created by permutations (n = 1,000). 331 individuals with IC/BPS compared to 6,516 controls.

### Families with multiple affected members

We examined families with multiple symptomatic family members and at least one asymptomatic family member to assess variants that segregated with symptoms (Table S4 – S5). Eight genes harbored rare variants present in affected (and absent in unaffected) members in at least two families. No genes were notable for previously identified Mendelian conditions associated with IC/BPS. *PIEZO2* (MIM: 613629) encodes a mechanically activated ion channel Piezo2 implicated in arthrogryposis (*92*). *PIEZO2* may mediate visceral hypersensitivity in irritable bowel syndrome via 5-HT release (*93*).

### Trio analyses

We examined 13 available trios from the BCH cohort with unaffected parents and an affected proband (Table S6) for *de novo* variants and compound heterozygosity. No genes were notable for previously identified Mendelian conditions associated with IC/BPS. One *de novo* variant was in *PLXNA4* (MIM: 604280), which has been referenced as a candidate for host response and wound healing in the bladder urothelium (*94*). Additional *de novo* variants were identified in *CS* (MIM: 118950), *HCN2* (MIM: 602781), *USP34* (MIM: 615295), and *GCNT4* (MIM: 616782). Inherited, compound heterozygous variants were identified in *LRP1* (MIM: 107770), *FRAS1* (MIM: 607830), and *SPEG* (MIM: 615950). The established phenotypes associated with these genes (Table S6) did not have phenotype overlap with IC/BPS. Multiple genes with *de novo* variants are intolerant to loss-of-function variant (Table S6) although all variants were missense variants. A gene set analysis of these genes did not yield a significant association (Table S2, Data S6).

### Copy number variant (CNV) analysis

We reviewed deletions detected using XHMM (*95*). We did not detect any deletions in genes included in our diagnostic analysis as well as genes of interest from our association studies. However, we identified one patient with a pathogenic CNV (Table S8) associated with chromosome 16p11.2 duplication syndrome (MIM: 614671) which has also been identified in renal hypodysplasia and congenital anomalies of the kidney and urinary tract (CAKUT) (*96, 97*). Deletions associated with autosomal recessive conditions were detected but no additional pathogenic variants in the affected gene were detected (Table S8).

## Discussion

We conducted a phenome-wide comorbidity analysis, exome-wide analysis of ultra-rare and rare damaging variants in IC/BPS encompassing collapsing-based gene burden tests, single variant analyses, gene-set and network-based pathway analyses, family-based studies and CNV diagnostic evaluation to identify genetic risk factors for IC/BPS. The unbiased studies uncovered new comorbidities and new biological pathways likely to be important in the etiology of IC/BPS.

Phenotype analyses using phecodes from eMERGE validated known associations while broadening the possibility of additional related phenotypes (Fig. 1). Confirmed associations included dysuria (*1, 2*), polyuria (*1, 2*), irritable bowel syndrome (*30, 98*), allergies (*41*), nausea (*42*), and myalgia (*30*). An association with GERD was also confirmed (*27, 40*). Many previously reported comorbid conditions for IC/BPS, including migraine and psychiatric disorders (anxiety disorders, depression, and mood disorders) were not significantly associated after correction. This is likely due to the small sample size and in no way negates the reality of these conditions in the IC/BPS population. Rather, it elevates the importance of some common but more under-recognized pain related conditions such as GERD and its downstream consequences including esophagitis, esophagus ulcers, and Barrett’s esophagus. The novel association with Barrett’s esophagus is significant due to its increased risk of cancer (*99*). Barrett’s and esophageal ulcer are likely downstream effects of the association between IC/BPS and GERD (*100*) reinforcing the necessity for multidisciplinary care for individuals with IC/BPS (*2*) and providing additional evidence for a common pathophysiology based on epithelial dysfunction (*40*). The association with osteoarthrosis/osteoarthritis is interesting due to the possibility of shared etiology; for example, Pentosan Polysulfate Sodium, an oral FDA-approved treatment for IC/BPS (*2, 101*), has been proposed for repurposing in osteoarthrosis/osteoarthritis (*102*). In addition, tiaprofenic acid, a non-steroidal drug for inflammatory pain, has been linked to chemical cystitis that can continue after discontinuing treatment (*103*).

Although to our knowledge, this is the largest genetic study of IC/BPS, none of the signals in the gene-based collapsing analyses and ExWAS reached exome-wide significance, reflecting the genetic heterogeneity of disease (Fig. 2, Fig. 4). Our study thus nominates multiple new candidate genes that will require confirmation in independent studies including some without current Online Mendelian Inheritance in Man (OMIM) disease associations such as *HTR7*, *MNT*, *CALB2*, *STX6*, and *TBC1D26* (Table 2) (*104*). *HTR7* encodes a 5-HT receptor implicated in visceral hypersensitivity in IBS (*49, 105*). Individuals with IC/BPS are at increased risk of having IBS (*24*). Another gene of interest is *DNAAF4* (also known as *DNAAF* and *DYX1C1*), which is linked to dyslexia susceptibility and ciliary dyskinesia (*48, 106*). Interestingly, a significant increase in deleterious variants in this and other cilia biogenesis genes was identified in a study of familial urinary bladder cancer (*107*). In one family with multiple affected family members (Table S5, S6), we identified a rare segregating variant in *PIEZO2*, encoding a mechanosensitive ion channel implicated in urethra/bladder control of micturition (*108*). The analysis of the small number of available trios (Table S6) yielded a *de novo* variant in *PLXNA4,* which has been linked to a new form of bladder epithelial cells and discussed in the context of IC/BPS (*94*). Importantly, we confirmed the association with *ATP2C1* and *ATP2A2*, which have been implicated in the desquamating skin disorders Hailey-Hailey and Darier disease (Table S2). *ATP2C1* and *ATP2A2* are expressed in the urothelium, suggesting the calcium transporter-related epithelial injury observed in these syndromes may also extend to the bladder urothelium and produce the symptoms of IC/BPS (*23, 40*).

In the setting of genetic heterogeneity, pathway-based tests can improve analytic power by detecting signals that are distributed across biologically interconnected genes (Table 2, Fig. 3). These analyses provided no support for previously implicated pathways such as urogenital development, pain perception, depression, and irritable bowel syndrome. However, the NHC based analysis yielded multiple new candidate genes and pathways (Table 3, Data S7). *SMAD3,* which is included in the most enriched gene cluster, has been implicated as a therapeutic target for interstitial cystitis (*109, 110*) and in constipation-associated neuropathic pain (*111*). Notably, additional genes within the cluster (*ANAPC7* [MIM: 606949], *CDC20* [MIM: 603618], and *SKIL* [MIM: 165340]) are linked to bladder cancer (*112–114*), and *FOXO1* is tied to urothelial fibrosis (*115*). Other top pathways included the positive regulation of MAPK cascade (Table 3) and the integrin/focal adhesion networks (Table 3, Data S7 Cluster 14). These pathways have been implicated in urothelial injury and bladder cancer progression. We found a significant association with the “small molecule transport” pathway (Fig. 3) which also includes *ATP2C1* and *ATP2A2* (*116*). These genes and pathways provide new avenues for investigation in larger human cohorts and model organisms.

There were several limitations of this study. Full phenotype data of the individuals with exome sequencing data were not always available preventing confirmation of clinical concordance with genotypic data. Our case size, while sizable for an IC/BPS study, is underpowered, reflecting the challenges associated with genetic studies of underdiagnosed diseases. We only investigated rare variant contributions to IC/BPS. Hence, an analysis of common variants may identify additional gene or pathways or support previously implicated pathways such as pain perception or depression. Finally, phenotype analyses using phecodes from eMERGE validated known associations while broadening the possibility of additional related phenotypes, but these analyses are vulnerable to the presence of an uncorrected confounding variable.

Overall, our study has several clinical and research implications. The phenotype associations encourage clinicians caring for IC/BPS patients to be vigilant about comorbid conditions including GERD, osteoarthrosis/osteoarthritis, Barrett’s esophagus and disorders of the epithelium. We did not find genetic evidence for some of the previously proposed pathogenic pathways in IC/BPS, but we identified predicted-damaging rare variants for monogenic forms of blistering skin diseases, and evidence for a common pathway for those genes.. Furthermore, this study supports the association of IC/BPS with several new pathways such as *PI3K*, cell cycle and integrin signaling as potential pathogenic mechanisms of disease. In addition, our study indicates that a larger sample size and an investigation of common variants would continue to better delineate the genetic architecture of IC/BPS.

## Materials and Methods

### Participants, Phenotyping, and Informed Consent

This study includes two main cohorts. (1) The “BCH cohort” was previously described (*23*). Inclusion criteria included IC/BPS diagnosis from physician, and symptoms of urinary urgency and pelvic, suprapubic and/or abdominal pain for at least 3 months in a 6-month period. Exclusion criteria evidence of a recent urinary tract infection (within the prior 3 months), structural urinary tract abnormalities, or cancers of the ladder, prostate, cervix, or uterus. Probands could have sporadic or familial IC/BPS. Additional phenotypes and comorbid conditions were extracted from available medical records and questionnaires. Probands could be either children or adults with an IC/BPS diagnosis and lived in the US or Canada. This cohort was enrolled following informed consent under an IRB-approved protocol at Boston Children’s Hospital (04-11-160).

(2) The “MaGIC cohort” included DNA from 100 randomly selected unrelated individuals from The Maryland Genetics of Interstitial Cystitis Study (MaGIC) (*36*). The MaGIC cohort consists of families with two or more blood relatives with IC/BPS. Documented inclusion criteria for this cohort included the following: 13 years of age or older and diagnosed with IC/BPS; having at least one relative with IC/BPS or similar urinary symptoms; were available for an interview by phone, mail, or website; and lived in the US or Canada. Individuals meeting the above criteria who had Parkinson’s disease, Multiple Sclerosis, Spina bifida, or another neurologic disease that preceded interstitial cystitis symptoms were excluded. Available additional phenotypes and comorbid conditions were obtained from sample metadata (*36*). These samples were received anonymized and covered under IRB-approved protocol at Columbia University Irving Medical Center (AAAS7948).

Combining the MaGIC and BCH cohorts created the “combined cohort”. We also analyzed 7 families with more than 1 affected family member and at least 1 unaffected family member and 13 trios with unaffected parents.

### Phenotype analysis

Phenotype data was obtained through eMERGE-III (*39*). To test the association between interstitial cystitis and other phenotypes phenome-wide, we first harmonized the coded diagnoses data by converting all available ICD-10-CM codes to the ICD-9-CM system. This approach was motivated by the facts that the great majority of data for eMERGE-III participants is already coded using the ICD-9 system; ICD-10 codes are more granular and thus, a reverse conversion is more prone to mapping errors; and the current PheWAS R library supports only ICD-9 codes (*38, 117*). The data set initially contained 102,739 individuals with 19,411 ICD9 codes. 5 codes were excluded (X999, NoD.x, NoI9, IMO0001, IMO0002) and individuals with unavailable or ambiguous sex were excluded. Subjects with ICD9 code 595.1 were considered individuals affected with IC/BPS (360 individuals). Individuals without ICD9 code 595.1. were considered unaffected (99,249 individuals). ICD9 codes were then transformed into 1,817 unique phecodes with descriptions using the mapCodesToPhecodes and addPhecodeInfo functions in the R PheWAS library (*118*). All individuals with ICD9 code 595.1 were assigned phecode ”chronic interstitial cystitis” (592.13). For all phecodes other than 595.1, case/control carrier status was determined using createPhenotypes function in the R PheWAS library using default settings including add.phecode.exclusions = T but modified min.code.count = 10 so that the case definition requires a minimum of 10 ICD-9 codes from the “case” grouping of each phecode.

The PheWAS function in the R PheWAS library was utilized to implement logistic regression. The package uses predefined “control” groups for each phecode. In total, all 1816 phecodes were tested using logistic regression with each phecode patient-control status as an outcome and chronic interstitial cystitis adjusted for sex, age, recruitment site, total ICD9 codes, and 9 ancestry principal components as a covariate. To establish significant disease associations in PheWAS, we set the Bonferroni-corrected statistical significance threshold at 2.8×10−05 (0.05 divided by 1816), correcting for 1816 independent phecodes tested. PheWAS plots were created using phewasManhattan function from the R PheWAS library.

### Next-Generation Sequencing Data Generation

Exome sequencing (ES) from the “BCH cohort” has been described previously (*23*). Samples were sent to the Broad Institute (Cambridge, MA) for Illumina-based short read ES. ES from the MaGIC cohort sent to Psomagen Inc. (Rockville, MD) for Illumina-based short read ES.

Exome sequencing (ES) of cases were transferred to the Institute for Genomic Medicine (IGM) at Columbia University Irving Medical Center (CUIMC). Controls were either sequenced at or transferred to IGM. Data were either ES (n = 10,696) and genome sequencing (GS, n = 1,285). ES and GS for more than 120,000 individuals have undergone harmonized alignment and variant calling as described previously (*119–121*). Our in-house Analysis Tool for Annotated Variants (ATAV) platform was used for variant filtering (*122*).

### Exome Sequencing and Genome Sequencing Data Generation

Multiple capture kits were used for exome sequencing and sequenced according to standard protocols with 150 bp paired-end reads. Genomes were sequenced according to standard protocols. Possible bias introduced by using ES from different capture kits (Table S9) and GS in collapsing is corrected during coverage (*43, 46*).

### Harmonized Alignment and Variant Calling

Individuals with IC/BPS (cases) and without (controls) were processed with the same bioinformatic pipeline for alignment and variant calling which has been used at the IGM to generate ES or GS for more than 120,000 individuals (*119–121*). DRAGEN (Edico Genome, San Diego, CA, USA) (*123*) was used to align reads to human reference GRCh37. Picard (Broad Institute, Boston, MA, USA) was used to mark duplicates (*124*). Genome Analysis Toolkit (GATK - Broad Institute, Boston, MA, USA) Best Practices recommendations v.3.6 was used for variant calling and annotated was completed with ClinEff (*125–127*). IGM’s in-house analysis tool for annotated variants (ATAV) (*122*) platform was used to add custom annotations including Genome Aggregation Database (gnomAD) v.2.1 frequencies (*128*), the probability of being loss-of-function (LoF) intolerant (pLI) (*129*), Transcript-inferred Pathogenicity (TraP) score (*89*), and loss-of-function observed/expected upper bound fraction (LOEUF) (*128*) scores and deciles. The collapsing workflow utilizes gene symbol matching between consensus coding sequence (CCDS release 20) (*130*). 18,852 CCDS v20 genes were called as part of the collapsing pipeline.

### Collapsing Analysis

Collapsing analysis is used to identify an association between cases status and rare-variant carrier status at the gene-level (*43–46*). We used the following steps: i) choose high-quality cases and selected controls enrolled broadly as “healthy family members”, “control mild neuropsychiatric disease” or “control”, and remove related individuals, ii) create clusters with cases and controls matched using geographic ancestry, iii) harmonize coverage separately in each cluster, iv) specify QV criteria to create collapsing models, v) assign indicator variable (0/1) to each case and control based on the absence/present of a QV in the gene, tested gene set, or variant, vi) test for association between case/control status and indicator variable using Cochran-Mantel-Haenszel (CMH) test, and vii) visualize results.

### Sample Quality Control (QC) and Removal of Related Individuals

Sequencing data from cases and controls passed the same quality control. Samples were only included if at least 90% of the consensus coding sequence (CCDS release 20)(*130*) covered at a minimum of 10x, less or equal 2% contamination levels according to VerifyBamID (*131*), and single nucleotide variants (SNVs) and indels overlapping the Single Nucleotide Polymorphism database (dbSNP) (*132*) at least 85% and 80%, respectively. Samples with discordance between self-declared and sequence-derived sex were excluded to prevent phenotype-genotype mismatch. KING was used to remove one of each pair of any related individuals with an inferred relationship of second-degree or closer while favoring inclusion of cases over controls and well-covered over poorly covered samples (*133*). 2 individuals were removed because unclear metadata-sample pairing, 1 individual’s sequencing data were removed from collapsing due to less than 90% of the CCDS region being covered by 10x (see Materials and Methods). This left 380 individuals with IC/BPS (352 females) for rare variant analysis who were compared to 11,981 unaffected individuals. After removing related individuals including additional duplicate samples, 348 individuals with IC/BPS (325 females) and 11,669 controls (6,596 females) remained with sequence data.

### Clustering

Clustering was performed as previously described (*43–45*). Appropriately matching case and controls for geographic ancestry attempts to account for population sub-structure (*134–136*). Principal component analysis on a set of pre-defined variants to capture population substructure (*137*). The Louvain method of community detection was utilized on the first six principal components to identify clusters reflecting the ancestry captured by the pre-defined variants mentioned above (*45, 138*). In addition, a geographic ancestry label was assigned to each sample using a pre-trained neural-network generated probability estimates for each of six groups (European, African, Latino, East Asian, South Asian and Middle Eastern). A 95% probability cut-off was used for the ancestry groups, and “Admixed” samples were those that did not reach 95% for any of the ancestry groups (Fig. S4). The neural-network assigned ancestries were used only to visually inspect the approximate composition of the clusters created by the Louvain method. We visually inspected the compositions of clusters using the Uniform Manifold Approximation and Projection (UMAP) to visualize overlap between cluster membership and predicted geographic ancestry (*139–141*). Clusters containing at least 15 cases and 15 controls were included in all analyses based on collapsing clusters (*44*). All clusters underwent coverage. Of the 348 individuals with IC/BPS that underwent clustering, 331 individuals were in included clusters (309 female). Of the 11,669 controls that underwent clustering, 6,516 were in included clusters (3,424 female).

### Coverage Harmonization

Because the included cases and controls have varied coverage throughout the coding region (exome vs. genome sequencing, multiple capture kits), we reduce bias introduced by unbalanced coverage between cases and controls using “coverage harmonization” (*46*). We have already screened samples to ensure that 90% of the consensus coding sequence (CCDS release 20) is covered at more than 10x. Only genomic sites covered more than 10x are included. We removed sites using a site-based pruning approach where the absolute difference in percentages of cases compared to controls with at least 10x coverage was greater than 7.0%. Coverage harmonization reduces the influence of coverage differences caused by different capture kits, inclusion of both ES and GS or sequencing depth in general (*44, 142, 143*). Coverage harmonization is performed on each cluster independently (Fig. S1).

For collapsing analyses, only sites where at least 80% of cases and 80% of controls had greater than 10x coverage included. For the regression ExWAS, only sites where at least 90% of cases and 90% of controls had greater than 10x coverage included.

### Variant-Based Collapsing Exome Wide Association Study (ExWAS)

We filtered out variants with allele count less than 10. All variants were covered at least 10x in 90% of cases and 90% of controls. All variants have a minor allele frequency < 1% in both gnomAD genome and exome. At a cluster level, the internal allele frequency was allowed to exceed 1% only by one allele. We extracted the number of cases/controls with and without a QV per gene and used the exact two-sided Cochran-Mantel-Haenszel (CMH) test to test for an enrichment of qualifying variants while controlling for cluster membership (*44, 45, 144–146*).

### Qualifying Variant Definition

Each gene-based collapsing model is defined by the inclusion criteria for a “qualifying variant” (QV) which are considered equivalent (*46, 147*). QV/Model parameters included external minor allele frequency (MAF), internal allele frequency, variant effect, and *in-silico filters* (Table S1). External frequency filters in gnomAD could be either “ultra-rare” (absent) or “flex” (MAF < 0.1%). For the “flex” models, MAF was filtered at a population specific level using gnomAD exomes. For gnomAD exomes, populations included afr, amr, asj, eas, sas, fin, and nfe. For gnomAD genomes, the MAF filter was applied to the full population. Internal allele frequencies were applied by cluster. For each cluster in the ultra-rare models, variants were excluded with an internal allele frequency greater than 0.05% applied to the combined case-control call set by cluster after excluding one allele (*148*). For flex models, the internal allele frequency filter was set at 1%.

For all variants in the ultra-rare and flex models, those with a proportion expression across transcripts (pext) value (when available) less than or equal to 50% the maximum pext value for that gene were removed as they are less likely to affect translated mRNA (*148*). All predicted loss-of-function (LOF) variants (stop gain, frameshift, splice acceptor, and splice donor variants) were filtered with Loss-Of-Function Transcript Effect Estimator (LOFTEE) to remove likely false-positive LOFs (*128*).

### Gene-Based Collapsing

We extracted the number of cases/controls with and without a QV per gene and used the exact two-sided Cochran-Mantel-Haenszel (CMH) test to test for an enrichment of qualifying variants while controlling for cluster membership (*44, 45, 144, 146*).

### Quantile-Quantile Plots and Genomic Inflation Factor **λ**

To visualize the distribution of statistical associations we created quantile-quantile (QQ) plots. The synonymous model was used as a negative control for gene-based collapsing (Figures S1 – S2). We defined a study-wide Bonferroni multiplicity-adjusted significance threshold of *p* < 5.4 x 10^-7^ (0.05 / [18,286 CCDS genes × 2 non-synonymous models + 55,607 variant sites in ExWAS]). No individual association met study-wide significance.

QQ plots for collapsing analyses were created for each model by plotting expected vs. observed *p-*values for each gene in the collapsing model. As described previously empirical (permutation-based) expected probability distributions were generated for each model independently (*44, 45*). Within each cluster, the case and control labels were randomly permuted while the rest of the gene by sample matrix was kept fixed. The number of newly randomly labeled cases/controls with and without a QV per gene were used for the CMH test to test for an association between case/control status and QV status while controlling for cluster membership (see Collapsing by Gene and Statistical Enrichment). An empirical distribution of 1,000 *p*-values for each gene was created by repeating this process 1,000 times. For each permutation the *p*-values were ordered. The mean of each rank-ordered estimate across the 1,000 permutations (i.e., the average 1st order statistic, the average 2nd order statistic, etc.) represented the empirical estimates of the expected ordered *p*-values. The genomic inflation factor λ based on the permutation-based expected *p*-values was estimated using a regression method as described previously (*149, 150*).

### Gene Set Collapsing Analysis

Analyzing sets of genes with known or suspected biological interactions can reveal disease-associated pathways by aggregated signal across related genes (*46, 150*). We tested the association between case/control status and harboring a variant in a gene-set by extracted the number of cases/controls with and without at least one QV among any of the genes in each of the gene-sets and used the exact two-sided CMH test (*43, 44, 144, 146*). We did not adjust for multiple comparisons.

### Analyzed Gene-Sets

We tested associations of case/control status with QV status in gene-sets in our combined European and Middle Eastern geographic ancestry cohorts. We tested associations in the following gene-sets:

1. 41 genes associated with congenital anomalies of the kidney and urinary tract (CAKUT) determined by the Invitae CAKUT panel (Test code: 434341)(Data S6) (*55*)
2. 31 genes associated with epithelial structure maintenance (GO:0010669) excluding *MUC2* (MIM: 158370) (Data S6)
3. 59 genes associated with morphogenesis of an epithelial sheet (GO: 0002011) excluding *CLASP2* (MIM: 605853), *MIR221* (MIM: 300568) (Data S6)
4. 5 genes previously implicated in Mendelian diagnoses among individuals with IC/BPS (*ATP2C1* [MIM: 604384], *DCAF8* [MIM: 615820], *SIX5* [MIM: 600963], *ENAM* [MIM: 606585], *ATP2A2* [MIM: 108740]) (Data S6) (*23*)
5. 79 genes associated with desquamating dermatologic disorders drawn from GeneDx dermatologic disorders panels (https://www.genedx.com/tests/Dermatology/dermatologic-disorders~c9693). Diseases included Darier Disease (MIM: 124200), Hailey-Hailey Disease (MIM: 169600), Epidermolysis Bullosa (Dystrophic) (MIM: 131850, 131750, 226600), Pachyonychia Congenita type 1 (MIM: 167200), Epidermolysis Bullosa Simplex (MIM: - PS131760), Epidermolytic Ichthyosis (MIM: PS607602, PS113800), Superficial Epidermolytic Ichthyosis (MIM: PS607602, PS113800), Bullous Ichthyosiform Erythroderma (MIM: PS607602, PS113800), Pachyonychia Congenita type 2 (MIM: PS167200), Epidermolytic palmoplantar keratoderma (MIM: PS144200), Epidermolysis Bullosa Junctional Type (MIM: PS226650), Generalized Atrophic Benign Epidermolysis Bullosa (GABEB), Herlitz Junctional Epidermolysis Bullosa, Mitis Junctional Epidermolysis Bullosa, Non-Herlitz Junctional Epidermolysis Bullosa, and Focal Dermal Hypoplasia (MIM; 305600) (Data S6).
6. GO chemical and genetic perturbations pathways involving both *ATP2C1* and *ATP2A2*. These gene sets include BLALOCK_ALZHEIMERS_DISEASE_DN (1237 genes), DIAZ_CHRONIC_MYELOGENOUS_LEUKEMIA_UP (1390 genes), ENK_UV_RESPONSE_KERATINOCYTE_DN (480 genes), LU_EZH2_TARGETS_DN (378 genes), MULLIGHAN_MLL_SIGNATURE_2_DN (274 genes) (Data S6). Gene sets were extracted from the Molecular Signatures Database (MSigDB) using msigdbr (*85, 151, 152*).
7. 2,257 genes expressed more than 50 transcripts per kilobase million (TPM) in the bladder per The Genotype-Tissue Expression (GTEx) database (Data S6) (*153*).
8. 44 genes with associations to pain intensity identified by GWAS (Data S6) (*75*).
9. 43 genes with associations to major depression identified by GWAS (Data S6) (*76*).
10. 7 genes with associations to irritable bowel syndrome by GWAS (Data S6) (*77*).
11. 11 genes with associations to anxiety by GWAS (Data S6) (*78*).
12. 51 genes associated with osteoarthroses/osteoarthritis (Data S6) (*81*).
13. 22 genes associated with bladder development (Data S6) (*56–70*).
14. 4 genes associated with IC/BPS in recent animal models (Data S6) (*71–74*).
15. 8 genes identified as harboring *de novo* variants or rare compound heterozygous pairings (Table S7, Data S6).

### Gene-Set Enrichment Analysis

We utilized pre-ranked gene-set enrichment analysis (GSEA) to identify biological pathways implicated in genes identified in the damaging ultra-rare variant collapsing model (Fig. 1, Data S3) (*85*). We used fast gene set enrichment analysis (FGSEA) to implement GSEA via the R package “fgsea” using hallmark gene sets from Molecular Signature Database (MSigDB) (*86, 154*). 459 genes with case enrichment and unadjusted p-values < 0.1 were provided for testing. We reported pathways significantly associated (P < 0.05) after multiple comparisons adjustment.

In addition, we utilized network-based heterogeneity clustering (NHC, updated February, 2024) to identify clusters of genes linked by biological proximity and protein-protein interactions (PPIs) (*82, 155*). NHC may be employed to analyzed small cohorts when multiple, but related, causative genes are suspected. Once gene clusters enriched in cases compared to controls are identified, NHC will perform pathway and ontology enrichment analyses. NHC uses 3 ancestry principal components. We used the ultra-rare damaging model drawn from the European ancestry cohort (see Clustering in Methods). From this cluster, we extracted a gene list per individual with and out harboring an ultra-rare damaging variant for both individuals with IC/BPS (case) and those without (control). The algorithm produces gene clusters based on gene/protein interconnectivity and then identifies gene networks from these clusters with case enrichment. Using the case enriched clusters, the algorithm assesses genset enrichment for MSigDB Hallmark (50 genesets), KEGG Pathway (805 genesets), Reactome Pathway (1670 genesets), Wiki Pathway (791 genesets), GO Biological Process (7375 genesets), GO Molecular Function (1753 genesets). Gene clusters significance threshold was p-value < 0.01. NHC uses adjusted p-value 1e-5 as the significance cutoff for the geneset enrichment analysis. The analysis was run with the following parameters: --edge 0.99 –hub 100 –merge 0.5, -- mode 2 –network Y –boost N.

### Family Enrichment Analysis

We analyzed 7 families that had multiple affected family members and at least one non-affected member to identify common genes or variants. Qualifying variants (QVs) were heterozygous variants in all affected family members and absent from all unaffected family members. QVs had to meet the following quality metrics: QUAL > 30, GQ ≥ 20, -- min-ad-alt 3. Only variants present less than 1% in gnomAD genome and gnomAD exome were considered.

We identified gene-variant effect pairs where a gene harboring a QV appeared in at least 2 families and the QV had the same effect (e.g., missense, synonymous, loss-of-function).

### Copy number variant detection from exome sequencing

Copy-number variant (CNV) calling was performed on whole-exome sequencing data using the eXome-Hidden Markov Model (XHMM) software package (63). The XHMM analysis followed the tutorial provided at https://zzz.bwh.harvard.edu/xhmm/tutorial.shtml, using the recommended default parameters. The analysis was run in batches for two different target interval files: IDTERPv1 and Agilent V6. Identified CNVs were annotated using AnnotSV version 3.4.2. (*156*). We filtered CNV calls for genomic disorders, defined as CNVs that had at least 70% overlap of their span with a set of known pathogenic CNVs’ coordinates as previously described (*157*).

### *De novo* analysis

Thirteen probands had unaffected parents available for trio analyses. Data were analyzed on the Codified Genomics platform (San Diego, CA). Variants were phased with parental sequences to identify *de novo* and compound heterozygous variants. Variants were filtered for genotype quality (GQL>L20) and read depth (>10x). Combined Annotation Dependent Depletion (CADD) was used for *in-silico* predictions of the functional effect of missense mutations and SpliceAI for presumed splice variants.

‘Qualifying variants’ were defined as those that satisfied each of the following criteria: i) de-novo ii) minor allele frequencyL<10 in gnomAD for *de novo* variants and <0.02 for compound heterozygous variants (http://gnomad.broadinstitute.org/), iii) missense or loss of function or frameshift or affecting a splice-site and iv) if missense, in silico predicted damaging by SIFT, Polyphen-2 or a CADD scoreL>L10. ACMG guidelines were followed to quantify the pathogenicity of qualifying variants (*158*).

## Data analysis and display

Unless otherwise noted in the methods, data analysis and visualization were performed with R (v.4.3.1) (*159*).

## Data Availability

All data produced in the present study are available upon reasonable request to the authors

## Funding

This publication was supported by the Centers for Disease Control and Prevention of the U.S. Department of Health and Human Services (HHS) as part of a financial assistance award totaling $2,400,000 with 60 percent funding from the CDC/HHS (U01DP006634-01). The contents are those of the author(s) and do not necessarily represent the official views of, nor are they endorsed by, the CDC/HHS or the U.S. Government. (CAB)

National Institutes of Health grant 1K08HG012374 (JEM)

NewYork-Presbyterian Samberg Scholar (JEM)

Thrasher Early Career Research Award (JEM)

NIH/NIDDK CAIRIBU Interactions Core U24-DK-127726 (CAB)

NICHD Boston Children’s Hospital Intellectual and Developmental Disabilities Research Center Molecular Genetics Core Facility U54HD090255

## Author contributions

Conceptualization: JEM, AGG, CAB

Data curation: JEM, AM, SKM, AK, MV, MW, AK, KK, CAB

Formal Analysis: JEM, AM, SKM, MV, AK

Funding acquisition: JEM, AGG, CAB

Investigation: JEM, AM, SKM, CAB

Methodology: JEM, AM, SKM, JB, NH, MV, AK, CAB

Project administration: JEM, MW, CAB

Visualization: JEM, AM, SKM

Resources: AGG, CAB

Software: JEM, AM, SKM

Supervision: AGG, CAB

Visualization: JEM, CAB

Writing—original draft: JEM, SKM, CAB

Writing—review & editing: JEM, SKM, AGG, CAB

## Data and materials availability

The Genotype-Tissue Expression (GTEx) Project was supported by the Common Fund of the Office of the Director of the National Institutes of Health, and by NCI, NHGRI, NHLBI, NIDA, NIMH, and NINDS. The data used for the analyses described in this manuscript were obtained from: the GTEx Portal on 5/4/2023.

