## Supplement for "Interstitial Cystitis: a phenotype and rare variant exome sequencing study"

Joshua E. Motelow *et al*.

**This PDF file includes:**

Figs. S1 to S4

Tables S1 to S9

Data S1 to S8 (legends)

**Other Supplementary Materials for this manuscript include the following:**

Data S1 to S8

Supplementary Text

None

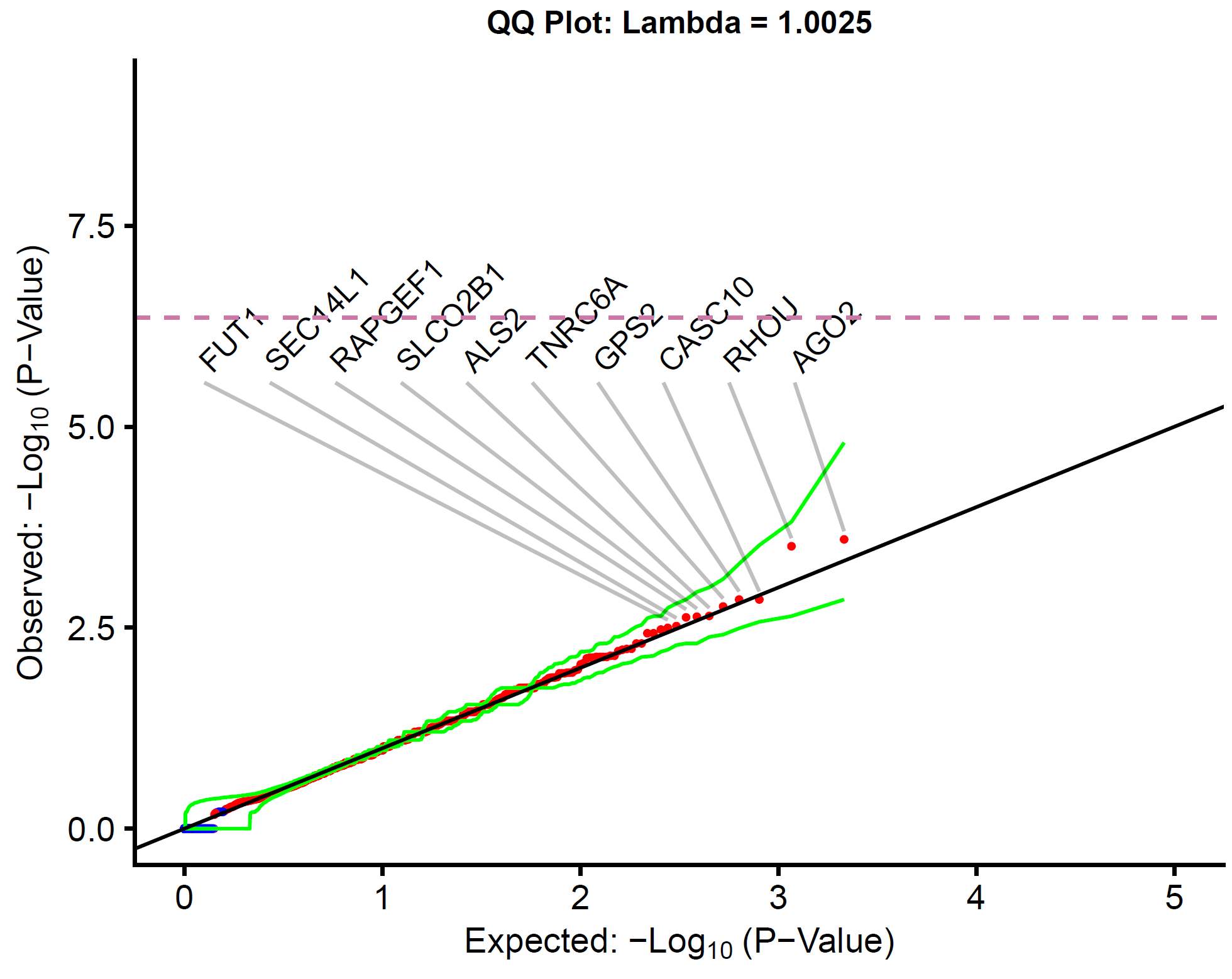

Fig. S1. Synonymous ultra-rare variant collapsing.

The quantile-quantile plots for the protein-coding genes with at least one case or control carrier of an ultra-rare synonymous variant. All variants are ultra-rare (i.e., allele frequency of less than 0.05% in internal case and control by cluster and absent in external reference cohorts, allele frequency < 0.00002 in internal database). P-values were generated from the exact two-sided Cochran-Mantel-Haenszel (CMH) test by gene by cluster to indicate a different carrier status of cases in comparison to controls. No gene achieved study-wide significance p < 5.4 x 10^-7^ after Bonferroni correction indicated by dashed line. Top ten case enriched genes are labeled. Red indicates case-enriched while blue indicates control enriched. The green lines represent the 95% confidence interval. Empiric confidence interval distributions created by permutations (n=1,000). 331 individuals with IC/BPS compared to 6,516 controls.

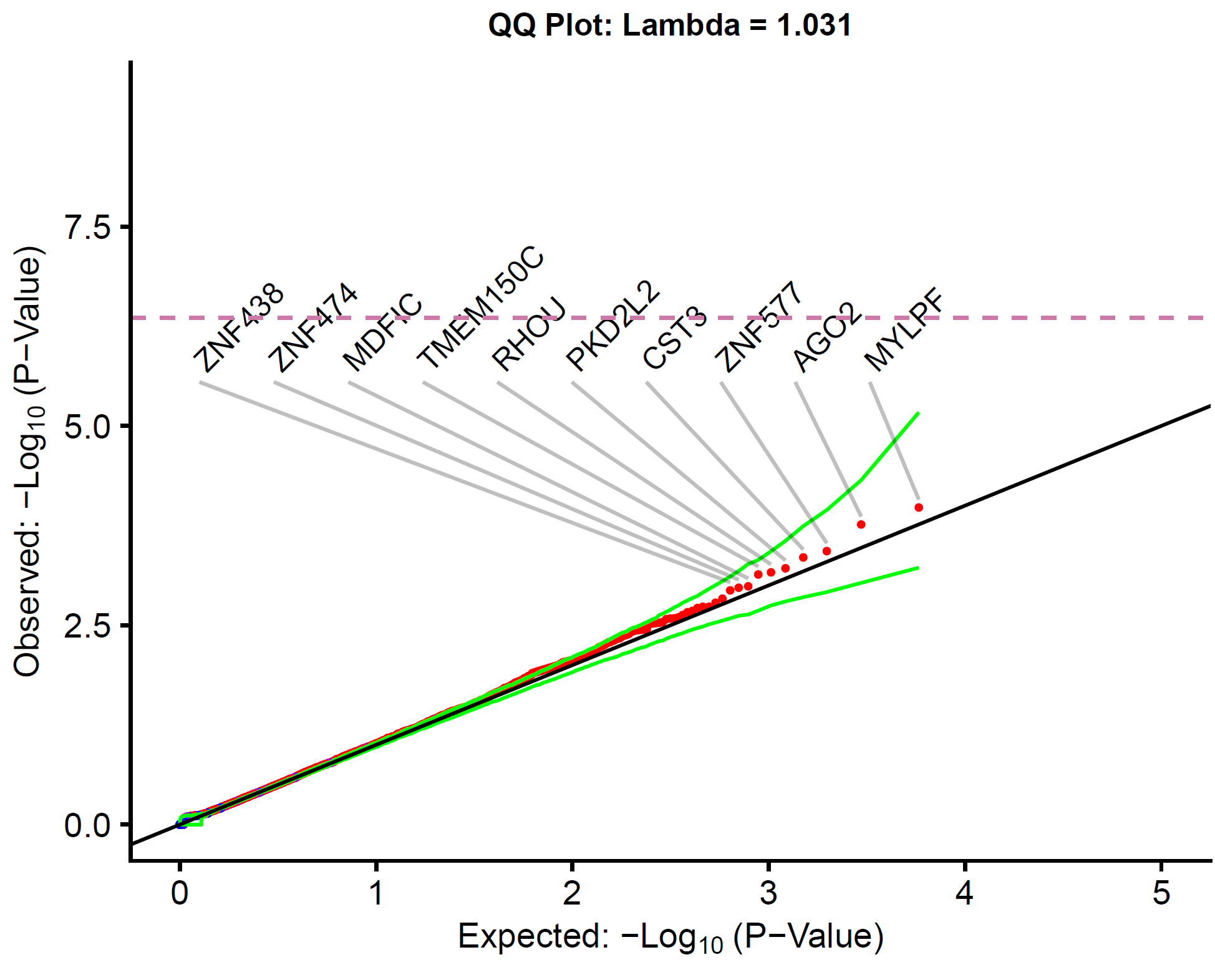

Fig. S2. Synonymous rare variant collapsing.

The quantile-quantile plots for the protein-coding genes with at least one case or control carrier of a rare synonymous variant. All variants are rare (i.e., allele frequency of less than 1% in internal case and control by cluster and less than 0.1% in external reference cohorts). P-values were generated from the exact two-sided Cochran-Mantel-Haenszel (CMH) test by gene by cluster to indicate a different carrier status of cases in comparison to controls. No gene achieved study-wide significance p < 5.4 x 10^-7^ after Bonferroni correction indicated by dashed line. Top ten case enriched genes are labeled. Red indicates case-enriched while blue indicates control enriched. The green lines represent the 95% confidence interval. Empiric confidence interval distributions created by permutations (n=1,000). 331 individuals with IC/BPS compared to 6,516 controls.

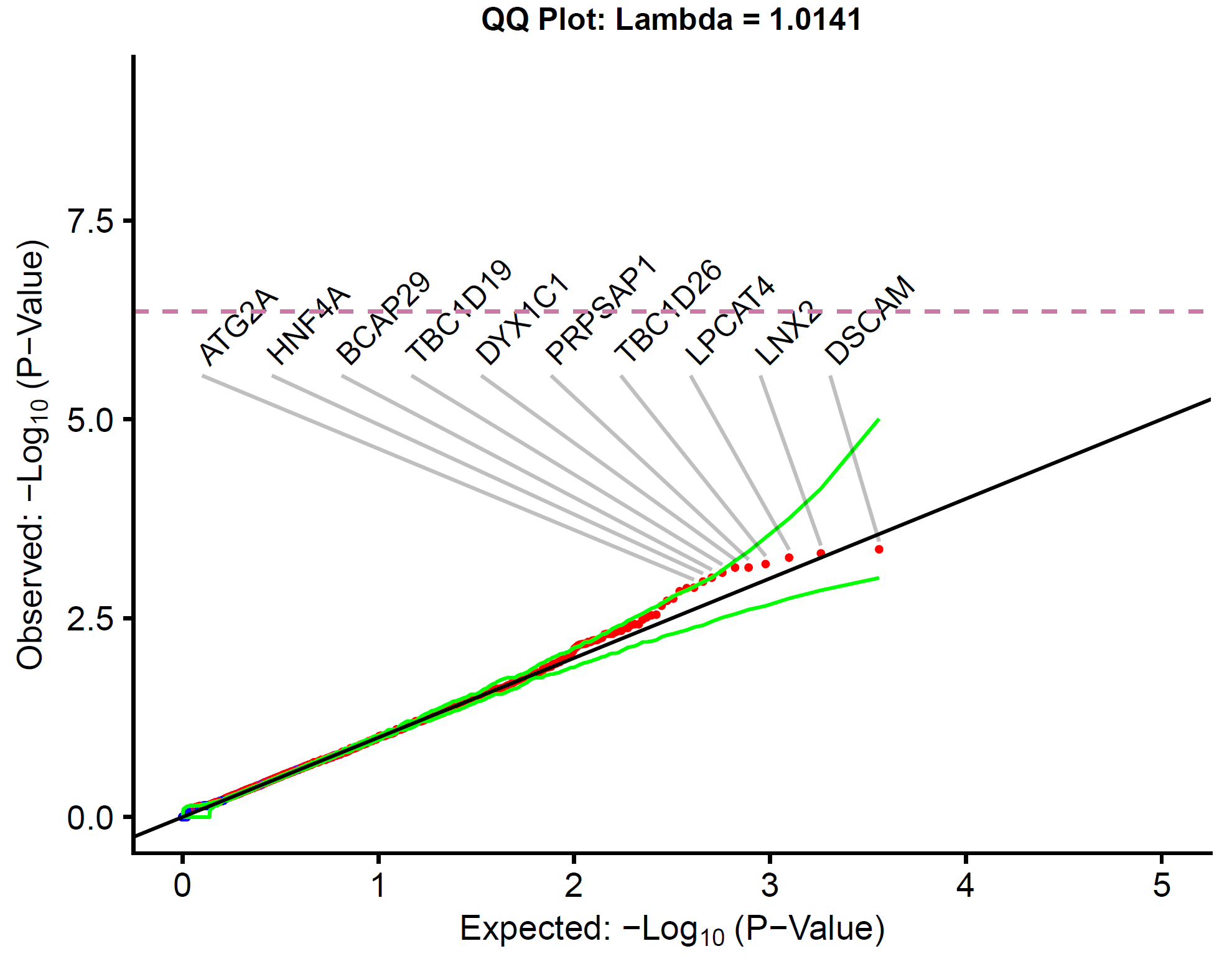

Fig. S3. Damaging rare variant collapsing.

The quantile-quantile plots for the protein-coding genes with at least one case or control carrier of a rare damaging variant. All variants are rare (i.e., allele frequency of less than 1% in internal case and control by cluster and less than 0.1% in external reference cohorts). P-values were generated from the exact two-sided Cochran-Mantel-Haenszel (CMH) test by gene by cluster to indicate a different carrier status of cases in comparison to controls. No gene achieved study-wide significance p < 5.4 x 10^-7^ after Bonferroni correction indicated by dashed line. Top ten case enriched genes are labeled. Red indicates case-enriched while blue indicates control enriched. The green lines represent the 95% confidence interval. Empiric confidence interval distributions created by permutations (n = 1,000). 331 individuals with IC/BPS compared to 6,516 controls. All missense REVEL > 0.5.

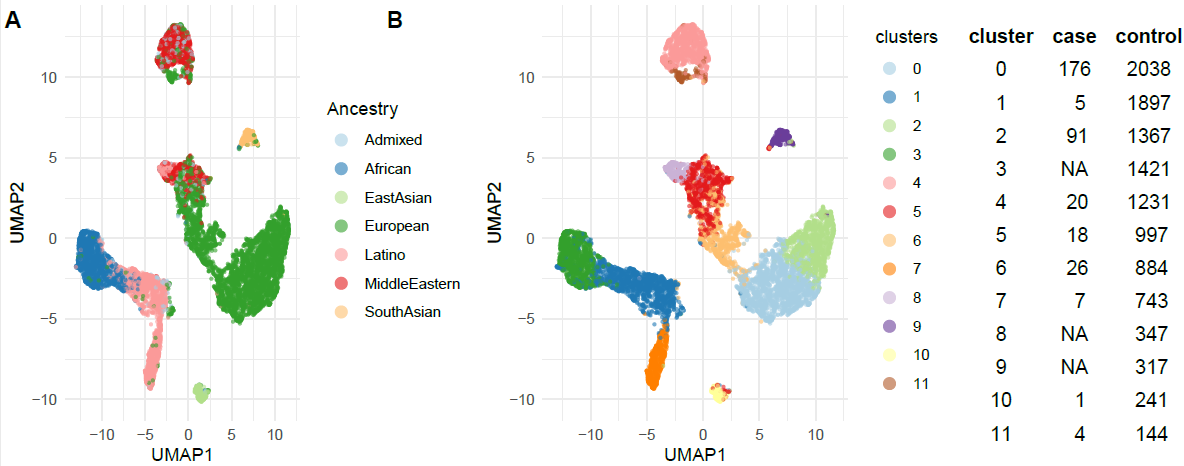

Fig. S4. UMAP of ancestry principle components.

UMAP and cluster assignments showing geographic ancestry of the case-control cohort. Only clusters with at least 15 cases and 15 controls (Clusters 0, 2, 4, 5, and 6) were included in the case-control analyses.

| Model Name | gnomAD Genome Max AF Global | gnomAD Exome Max AF Population Specific | Included Effects |
| --- | --- | --- | --- |
| Ultra-Rare Synonymous | Absent | Absent | Synonymous |
| Flex Synonymous | 0.05% | Population specific 0.05% | Synonymous |
| Ultra-Rare Damaging | Absent | Absent | LOF + Missense |
| Flex Damaging | 0.1% | Population specific 0.1% | LOF + Missense |
| ExWAS – Collapsing | 1% | 1% | Inclusive |
| ExWAS – European Only | 1% | 1% | Inclusive |

Table S1. Collapsing models

Summary of collapsing models and qualifying variant definitions. LOF = predicted loss-of-function effect. LOF effects include stop gained, frameshift, splice acceptor, and splice donor variants. Inclusive = exon loss variant, frameshift variant, rare amino acid variant, stop gained, start lost, stop lost, splice acceptor variant, splice donor variant, gene fusion, bidirectional gene fusion, 3 prime UTR truncation + exon loss variant, 5 prime UTR truncation + exon loss variant, coding sequence variant, disruptive inframe deletion, disruptive inframe insertion, conservative inframe deletion, conservative inframe insertion, missense variant + splice region variant, missense variant, splice region variant, 5 prime UTR premature start codon gain variant, initiator codon variant, initiator codon variant + non canonical start codon, splice region variant + synonymous variant, splice region variant, start retained, stop retained variant, synonymous variant. In “Damaging” models, LOFTEE and REVEL > 0.5 applied. Populations in gnomAD exome include afr,amr,asj,eas,sas,fin,nfe.

| Gene-set/Variant-ID and Cohort | OR | CI Low | CI High | Case with QV | Case without QV | Ctrl with QV | Ctrl  without QV | P Value |
| --- | --- | --- | --- | --- | --- | --- | --- | --- |
| IC Genes – Combined cohort | 7.4 | 1.6 | 26.4 | 4 | 327 | 11 | 6505 | 0.0052 |
| IC Genes –  BCH cohort | 5.3 | 0.6 | 25.7 | 2 | 237 | 11 | 6505 | 0.071 |
| IC Genes – MaGIC cohort | 13.5 | 1.4 | 68.2 | 2 | 90 | 11 | 6505 | 0.014 |
| IC Genes – IBS cohort | 23.5 | 2.4 | 119.1 | 2 | 52 | 11 | 6505 | 0.0049 |
| IC Genes – Anxiety cohort | 29.2 | 3.0 | 146.6 | 2 | 39 | 11 | 6505 | 0.0032 |
| IC Genes – Migraine cohort | 16.8 | 1.7 | 85.7 | 2 | 75 | 11 | 6505 | 0.0093 |
| IC Genes - Anxiety + IBS + Migraine cohort | 17.6 | 3.0 | 73.7 | 3 | 103 | 11 | 6505 | 0.0014 |
| Derm Desqu Dx Genes – Combined cohort | 1.5 | 0.8 | 2.4 | 18 | 313 | 249 | 6267 | 0.14 |
| Epi Morph – Combined cohort | 1.3 | 0.6 | 2.5 | 9 | 322 | 156 | 6360 | 0.44 |
| Epi Struct – Combined cohort | 1.0 | 0.2 | 3.1 | 3 | 328 | 58 | 6458 | 1 |
| Kidney Genes – Combined cohort | 1.0 | 0.8 | 1.3 | 81 | 250 | 1581 | 4935 | 0.95 |
| CAKUT Genes – Combined cohort | 1.1 | 0.5 | 2.4 | 8 | 323 | 138 | 6378 | 0.69 |
| Bladder Expressed Genes – Combined cohort | 1.0 | 0.8 | 1.2 | 127 | 204 | 2521 | 3995 | 0.86 |
| IC top exwas hits – Combined cohort | 0.9 | 0.3 | 2.0 | 6 | 325 | 133 | 6383 | 1 |
| MULLIGHAN – Combined cohort | 0.8 | 0.5 | 1.2 | 24 | 307 | 608 | 5908 | 0.24 |
| LU EZH2 – Combined cohort | 1.1 | 0.7 | 1.5 | 38 | 293 | 691 | 5825 | 0.71 |
| ENK UV – Combined cohort | 0.9 | 0.6 | 1.3 | 35 | 296 | 750 | 5766 | 0.54 |
| DIAZ CHRONIC – Combined cohort | 0.8 | 0.6 | 1.0 | 81 | 250 | 1934 | 4582 | 0.04 |
| BLALOCK ALZHEIMERS – Combined cohort | 0.8 | 0.6 | 1.0 | 71 | 260 | 1712 | 4804 | 0.10 |
| IC de novo – Combined cohort | 0.8 | 0.0 | 4.8 | 1 | 330 | 31 | 6485 | 1 |
| Pain GWAS – Combined cohort | 0.6 | 0.1 | 1.9 | 3 | 328 | 99 | 6417 | 0.63 |
| Major Depression GWAS – Combined cohort | 0.5 | 0.1 | 2.1 | 2 | 329 | 67 | 6449 | 0.58 |
| IBS GWAS – Combined cohort | 0 | 0 | 9.3 | 0 | 331 | 9 | 6507 | 1 |
| Anxiety GWAS – Combined cohort | 0.5 | 0 | 3.2 | 1 | 330 | 37 | 6479 | 1 |
| IC/BPS Animal Genes – Combined cohort | 0.9 | 0.02 | 6.1 | 1 | 330 | 18 | 6498 | 1 |
| Bladder Development Genes – Combined cohort | 0.6 | 0.01 | 3.7 | 1 | 330 | 36 | 6480 | 1 |

Table S2. Gene set analyses

Gene set burden testing with biologically informed gene sets. We used the ultra-rare damaging model for all analyses (see Methods). Cohort descriptions can be found in Methods and Results. Pooled odds ratio (ORs) and 95% confidence intervals, and unadjusted *P* values were generated from the exact 2-sided Cochran-Mantel-Haenszel test.

| Variant | P Value | Estimate | Conf Low | Conf High | Qual Case | Unqual Case | Qualified Ctrl | Unqualified Ctrl |
| --- | --- | --- | --- | --- | --- | --- | --- | --- |
| '1-48708137-C-A' | 6.6E-06 | 16.2 | 4.8 | 54.1 | 7 | 324 | 8 | 6508 |
| '17-7483627-A-G' | 2.0E-05 | 38.6 | 7.7 | 168.1 | 4 | 327 | 7 | 6509 |
| '2-242757938-G-A' | 2.2E-05 | 37.6 | 7.5 | 159.3 | 4 | 327 | 8 | 6508 |
| '19-17671248-C-G' | 7.0E-05 | 32.0 | 5.7 | 157.6 | 4 | 327 | 6 | 6510 |
| '12-101017586-G-A' | 7.2E-05 | 12.8 | 3.6 | 43.8 | 6 | 325 | 8 | 6508 |
| '3-1269658-G-A' | 0.00010 | 8.3 | 2.9 | 21.3 | 7 | 324 | 21 | 6495 |
| '13-95271496-G-A' | 0.00012 | 17.6 | 3.8 | 81.5 | 5 | 326 | 5 | 6511 |
| '7-148801038-A-G' | 0.00017 | 6.5 | 2.4 | 15.8 | 8 | 323 | 21 | 6495 |
| '14-89747304-G-A' | 0.00019 | 15.6 | 3.5 | 70.2 | 5 | 326 | 5 | 6511 |
| '15-91474511-T-TGTC' | 0.00021 | 7.7 | 2.6 | 20.8 | 7 | 324 | 14 | 6502 |

Table S3. Cochran–Mantel–Haenszel exome wide association study (ExWAS) top associations

Top 10 associations combining European and Middle Eastern ancestry ExWAS.

| Family | Unaffected | Affected including Proband |
| --- | --- | --- |
| A | 4 | 2 |
| B | 1 | 2 |
| C | 2 | 3 |
| D | 4 | 2 |
| E | 1 | 2 |
| F | 2 | 2 |
| G | 1 | 3 |

Table S4. Family composition for segregation analysis

Composition of families with multiple affected family members and at least one unaffected family member

| Family | Gene Name | Effect | GT | Position | HGVS_c | HGVS_p |
| --- | --- | --- | --- | --- | --- | --- |
| A | *ABCC11* | missense_variant | het | 16-48232179-T-C | c.2090A>G | p.Glu697Gly |
| C | *ABCC11* | missense_variant | het | 16-48261787-G-A | c.325C>T | p.Arg109Trp |
| B | *AGXT* | missense_variant | het | 2-241808753-G-A | c.332G>A | p.Arg111Gln |
| F | *AGXT* | missense_variant | het | 2-241808308-C-A | c.26C>A | p.Thr9Asn |
| A | *COL4A4* | missense_variant | het | 2-228004920-C-T | c.149G>A | p.Arg50Lys |
| A | *COL4A4* | splice_region_variant | het | 2-228012304-T-C | c.-101-4A>G | *NA* |
| G | *COL4A4* | missense_variant | het | 2-227964372-G-C | c.1063C>G | p.Pro355Ala |
| A | *ESPNL* | missense_variant | het | 2-239009082-G-A | c.22G>A | p.Val8Met |
| B | *ESPNL* | missense_variant | het | 2-239039829-G-A | c.2474G>A | p.Arg825Gln |
| A | *PATL1* | missense_variant | het | 11-59423150-G-A | c.877C>T | p.Leu293Phe |
| G | *PATL1* | missense_variant | het | 11-59423440-C-G | c.802G>C | p.Gly268Arg |
| B | *PIEZO2* | missense_variant | het | 18-10672692-G-A | c.53C>T | p.Ala18Val |
| C | *PIEZO2* | missense_variant | het | 18-10705436-C-T | c.5558G>A | p.Arg1853His |
| B | *RERE* | missense_variant | het | 1-8616562-C-T | c.697G>A | p.Val233Ile |
| C | *RERE* | missense_variant | het | 1-8420188-C-T | c.3379G>A | p.Ala1127Thr |
| A | *SPATC1* | missense_variant | het | 8-145086777-C-T | c.94C>T | p.Arg32Trp |
| B | *SPATC1* | missense_variant | het | 8-145101720-G-T | c.1639G>T | p.Gly547Cys |

Table S5. Family analysis gene-effect combinations

Gene-variant effect pairs where a gene harboring a QV appeared in at least 2 families and the QV had the same effect non-synonymous effect. Family structure can be found in Table S4.

| ID | *de novo* variant (hg19) | Compound heterozygous variant (hg19) | CADD/  spliceAI | ACMG criteria | pLI/LOEUF | Disease association |
| --- | --- | --- | --- | --- | --- | --- |
| Trio 1 | *HCN2* chr19:603948A>G NM_001194.4:c.1037A>G p.Tyr346Cys exon 2 | no QV | 24.5 | VUS/LP: PM2 Moderate PP3 Moderate PP2 Supporting | 0.18/0.63 | MIM: 602477 |
| Trio 2 | *PLXNA4* chr7:131844258C>T NM_020911.2:c.4634G>A p.Arg1545Gln exon 25 | no QV | 33 | VUS/LP: PM2 Moderate PP2 Supporting | 1/0.29 | N/A |
| Trio 3 | *CS* chr12:56676232TG>CA NM_004077.3:c.559_560inv p.Gln187Trp exon 6 | no QV | NA | VUS/LP: PM2 Moderate PP2 Supporting | 1/ 0.36 | N/A |
| Trio 4 | *GCNT4* chr5:74325327A>G NM_001366737.1:c.536T>C p.Leu179Ser exon 4 | no QV | 26.7 | VUS/LB: PM2 Moderate | 0/0.8 | N/A |
| Trio 5 | *USP34* chr2:61475663T>G NM_014709.4:c.6377A>C p.Lys2126Thr exon 49 | no QV | 26.1 | VUS/LB: PM2 Moderate | 1/0.15 | N/A |
| Trio 6 | no QV | *LRP1* chr12:57569339G>A NM_002332.3:c.3644G>A p.Gly1215Glu exon 23 | 25.8 | LB: PP2 Supporting BS2 Supporting BP6 Strong | 1/.0.12 | MIM: 107770 |
|  |  | *LRP1* chr12:57588369G>A NM_002332.3:c.8078G>A p.Arg2693His exon 50 | 26.9 | VUS: PM2 Moderate PP2 Supporting |  |  |
| Trio 7 | no QV | *FRAS1* chr4:79353587C>G NM_025074.7:c.5046C>G p.Asp1682Glu exon 38 | 18.29 | B: BS1 Strong BS2 Strong BP6 Strong | 0/0.73 | MIM: 219000 |
|  |  | *FRAS1* chr4:79308659C>T NM_025074.7:c.3779C>T p.Pro1260Leu exon 29 | 25.8 | VUS: PM2 Moderate |  |  |
| Trio 8 | no QV | *SPEG* chr2:220353527AGGTAC>TGGTAA NM_005876.5:c.8054_8059delinsTGGTAA p.Glu2685_Pro2687delinsValValThr exon 34 | NA | VUS: PM2 Moderate | 0/0.51 | MIM: 615959 |
|  |  | *SPEG* chr2:220342509T>G NM_005876.5:c.4826+2T>G | 28.3 / 0.55 acceptor loss, 0.98 donor loss | VUS/LP: PVS1 Very Strong |  |  |
| Trio 9 | no QV | no QV |  |  |  |  |
| Trio 10 | no QV | no QV |  |  |  |  |
| Trio 11 | no QV | no QV |  |  |  |  |
| Trio 12 | no QV | no QV |  |  |  |  |
| Trio 13 | no QV | no QV |  |  |  |  |

Table S6. Trio analysis for *de novo* and compound heterozygous SNV/Indel variants

*De novo* variants and potentially paired compound heterozygous damaging variants identified in 13 trios.

| SYNDROME_NAME | SYNDROME_INTERVAL | XHMM_INTERVAL | KB |
| --- | --- | --- | --- |
| 16p11.2_duplication_Autism | chr16:29548679-30098679 | 16:29675050-30199897 | 524.85 |

Table S7. Autosomal dominant Mendelian disorder-associated copy number variants (CNVs) in individuals with IC/BPS

Duplication detected by CNV pipeline associated with autosomal dominant disease. Genes affected include *ALDOA, ASPHD1, C16orf54, C16orf92, CDIPT, CORO1A, DOC2A, GDPD3, HIRIP3, INO80E, KCTD13, KIF22, LOC101928595, MAPK3, MAZ, MVP, PAGR1, PPP4C, PRRT2, QPRT, SEZ6L2, SPN, TAOK2, TBX6, TMEM219, YPEL3, ZG16*. No other CNVs associated with autosomal dominant diseases were identified.

| INTERVAL | KB | CytoBand | Gene | GenCC disease | OMIM phenotype | ACMG class |
| --- | --- | --- | --- | --- | --- | --- |
| 15:43888606-43941032 | 52.43 | q15.3 | STRC | autosomal recessive nonsyndromic hearing loss 16;hearing loss, autosomal recessive;nonsyndromic genetic hearing loss | Deafness, AR 16, 603720 (3) AR (MIM: 606440) | full=5 |
| 15:43903117-43991287 | 88.17 | q15.3 | STRC | autosomal recessive nonsyndromic hearing loss 16;hearing loss, autosomal recessive;nonsyndromic genetic hearing loss | Deafness, AR 16, 603720 (3) AR (MIM: 606440) | full=5 |
| 16:5053403-5123257 | 69.86 | p13.3 | ALG1 | ALG1-CDG | Congenital disorder of glycosylation, type Ik, 608540 (3) AR (MIM: 605907) | full=4 |
| 1:161514491-161600992 | 86.5 | q23.3 | FCGR3A | autosomal recessive primary immunodeficiency with defective spontaneous natural killer cell cytotoxicity | Immunodeficiency 20, 615707 (3) AR (MIM: 146740) | full=4 |
| 1:161514491-161600992 | 86.5 | q23.3 | FCGR3A | autosomal recessive primary immunodeficiency with defective spontaneous natural killer cell cytotoxicity | Immunodeficiency 20, 615707 (3) AR (MIM: 146740) | full=4 |
| 1:214787098-214837137 | 50.04 | q41 | CENPF | Stromme syndrome | Stromme syndrome, 243605 (3) AR (MIM: 600236) | full=4 |
| 2:110873265-110962545 | 89.28 | q13 | NPHP1 | Bardet-Biedl syndrome;Joubert syndrome with renal defect;Senior-Loken syndrome;nephronophthisis 1 | Joubert syndrome 4, 609583 (3) AR;Nephronophthisis 1, juvenile, 256100 (3) AR;Senior-Loken syndrome-1, 266900 (3) AR (MIM: 607100) | full=5 |
| 9:98638288-99007708 | 369.42 | q22.32 | ERCC6L2 | pancytopenia-developmental delay syndrome | Bone marrow failure syndrome 2, 615715 (3) AR (MIM: 615667) | full=4 |
| 9:98638288-99007708 | 369.42 | q22.32 | HSD17B3 | 46,XY disorder of sex development due to 17-beta-hydroxysteroid dehydrogenase 3 deficiency | Pseudohermaphroditism, male, with gynecomastia, 264300 (3) AR (MIM: 605573) | full=4 |

Table S8. Autosomal recessive Mendelian disorder-associated CNV variants in individuals with IC/BPS

Copy number variants detected in probands associated with recessive conditions. Only deletions were detected. The CNV pipeline is unable to determine if homozygous or heterozygous, but no paired pathogenic variant detected. The probability of being loss-of-function (LoF) intolerant (pLI) and loss-of-function observed/expected upper bound fraction (LOEUF) scores were extracted from the gnomAD browser v4.1.

| Capture Kit | Control | Case |
| --- | --- | --- |
| 65MB | 540 | NA |
| AgilentCRE | 2 | NA |
| AgilentV4 | 110 | NA |
| AgilentV5 | 12 | NA |
| AgilentV5UTR | 99 | NA |
| AgilentV6 | 2 | 250 |
| Agilentv5 | 4 | NA |
| Genome | 442 | NA |
| IDTERPv1 | 3304 | NA |
| IDTERPv2 | 28 | 98 |
| MedExome | 9 | NA |
| Roche | 7037 | NA |
| RocheV2 | 38 | NA |

Table S9. Capture kits

Distribution of exome capture kits and genome sequencing.

Data S1. eMERGE Comorbidity analysis

See Fig. 1 legend for details.

Data S2. Summary statistics for synonymous ultra-rare variant collapsing analysis

Data S3. Summary statistics for synonymous flex rare variant collapsing analysis

Data S4. Summary statistics for damaging ultra-rare variant collapsing analysis

Data S5. Summary statistics for damaging flex variant collapsing analysis

For Data S2 – S5, summary statistics by gene for collapsing analysis indicated in the table title. See Table S1 for collapsing model and qualifying variant parameters.

Data S6. Gene lists for gene set collapsing analysis

See legend for Table S2 and Methods for details

Data S7. Network-based heterogeneity clustering associations

See legend for Table 3 and methods for details.

Data S8. Summary statistics for exome wide association study (ExWAS) analysis

Summary statistics by gene for ExWAS. See legend for Fig. 4 for details.
